# Gut permeability and cognitive decline: A pilot investigation in the Northern Manhattan Study

**DOI:** 10.1101/2020.09.15.20195362

**Authors:** Tatjana Rundek, Sabita Roy, Mady Hornig, Ying Kuen Cheung, Hannah Gardener, Janet DeRosa, Bonnie Levin, Clinton B. Wright, Victor A. Del Brutto, Mitchell SV Elkind, Ralph L. Sacco

## Abstract

**Background:** Gut microbiota may impact cognitive function and decline, though data is limited. This pilot study examines the associations between gut dysbiosis products, plasma lipopolysaccharide (LPS) and soluble CD14 (sCD14), with cognitive decline and immune molecule activation among 40 participants in the longitudinal population-based Northern Manhattan Study.

**Methods:** We selected stroke- and dementia-free participants at baseline with high activation levels of core components of the immune signaling pathways underlying microbiota metabolite-cognitive associations (IL-1, IL-17, TNF). Participants were followed with up to three complete neuropsychological assessments.

**Results:** Elevated sCD14 was associated with high levels of IL-1 (p<0.05), whereas in samples where only IL-17 and TNF were increased, LPS and sCD14 levels were not elevated. LPS was associated with decline in global cognitive performance over 2-3 assessments (adjusted β=-0.023 per SD per year, 95%CI:-0.036, −0.010). The association between sCD14 and cognitive decline was marginal (adjusted β=-0.018 per SD per year, 95%CI:-0.040, 0.004).

**Conclusions:** These preliminary data support the hypothesis that gut dysbiosis leads to systemic and neuro-inflammation, and subsequently cognitive decline. Further large targeted and untargeted gut microbiota-derived metabolomic studies are needed.

---

The human gut is home to trillions of microbes that participate in a lifelong symbiotic relationship with their hosts and perform essential regulatory functions for health ranging from regulating nutrition and metabolism to the immune system(1). Gut microbes impact health in part by metabolizing dietary and host-derived substrates, thereby generating biologically active compounds including signaling compounds, biological precursors, and toxins(2). Recent studies implicate gut microbiota in the regulation of cognitive functions through a ‘gut-microbiome-brain axis’(3) that has a particular relevance in the elderly because it contributes to chronic low-grade inflammation and modulates age-related changes in innate immunity and cognitive functioning(4). Gut dysbiosis, disrupted intestinal integrity, and high serum levels of gut microbiota-derived metabolites have been linked to stroke and CVD(5) as well as to cognition, mild cognitive impairment and Alzheimer’s Disease(6).

Gut dysbiosis disrupts gut barrier function and causes translocation to the circulation of bacterial products such as lipopolysaccharide (LPS), which leads to monocyte activation with release of proinflammatory markers including soluble CD14 (sCD14)(7). Our pilot study examines the relationship of LPS and sCD14 plasma levels with cognitive decline and levels of activation in the selected immune molecules, Interleukin-1 (IL-1), IL-17, and tumor necrosis factor (TNF) that may be activated by gut dysbiosis products. We hypothesized that increased plasma levels of LPS and sCD14 – markers of gut dysbiosis and compromised intestinal integrity – would be associated with increased circulating pro-inflammatory immune molecules and with cognitive decline, specifically because cognitive decline might be driven by inflammation.

## Methods

From the prospective community-based Northern Manhattan Study (NOMAS), we selected 40 healthy stroke- and dementia-free participants with high levels of activation of IL-1, IL-17 and TNF, which are considered to be ‘core’ molecules of the immune signaling pathways most commonly proposed as either mediators or modulators of microbiota metabolite-cognitive associations. They were selected from a 60 immune marker panel and defined by their levels >75th percentile to address the centrality of these core molecules in terms of their known effects in the intestinal tract and interactions with immune cells that influence microbiota, and their high degree of correlation with the other immune molecules in the same pathway. We then created seven non-overlapping groups (abscissa of the graph) and a control group, with each group defined by a combination of molecules from the 3 pathways with levels above 2 standard deviation (SD) above the group mean. The final study sample of 40 subjects was formed by a random selection of 5 individuals meeting criteria for each group.

NOMAS has been described in prior publications, including details on neurocognitive battery(8). We used the composite cognitive score created from four cognitive domains (executive function, memory, language, and processing speed) at baseline and at up to two follow-up visits (5 participants had only baseline;16 had one follow-up visit; 19 had two follow-up visits).

Plasma LPS and sCD14 levels were analyzed using enzyme-linked immunosorbent assay (R&D Systems, Minneapolis) as previously described(9). Immune molecules (IL-1, IL17, TNF) were measured using a magnetic, bead-based 60-plex immunoassay (Affymetrix/eBioscience, Santa Clara)(10).

We calculated means±SD of LPS and sCD14 in plasma samples in the activated immune molecule groups defined above. The association between LPS and sCD14 was assessed by a linear regression. Parametric statistical differences between the two immune molecules groups were established with a t-test, and non-parametric differences between these groups were established with a Mann-Whitney U-test. Generalized estimating equations (GEE) were used to estimate the association of LPS and sCD14 with longitudinal cognitive scores adjusting for age, sex, race/ethnicity, and education. Interaction terms between visit time and the LPS and sCD14 was introduced in the GEE model, and associations between baseline cognitive scores and the plasma levels were evaluated as main effects in the model.

## Results

In our pilot sample (N=40, mean age=65+/-9years, 68% women, 68% Hispanic, 13% non-Hispanic Black, 20% non-Hispanic white), LPS ranged from 79-224 (mean=176+/-33) pg/ml and sCD14 from 1200-2400 (mean=1597+/-310) ng/ml. For every pg/ml increase in LPS, sCD14 increased by 2.40 ng/ml (SE=1.48, p=0.11).

We observed elevated levels of sCD14 (p<0.05) only in groups with high IL-1 levels (above 2SD; Figure 1A). Conversely, in samples where only IL-17 and TNF were increased, and not IL-1, LPS and sCD14 levels were not significantly elevated (Figure 1B).

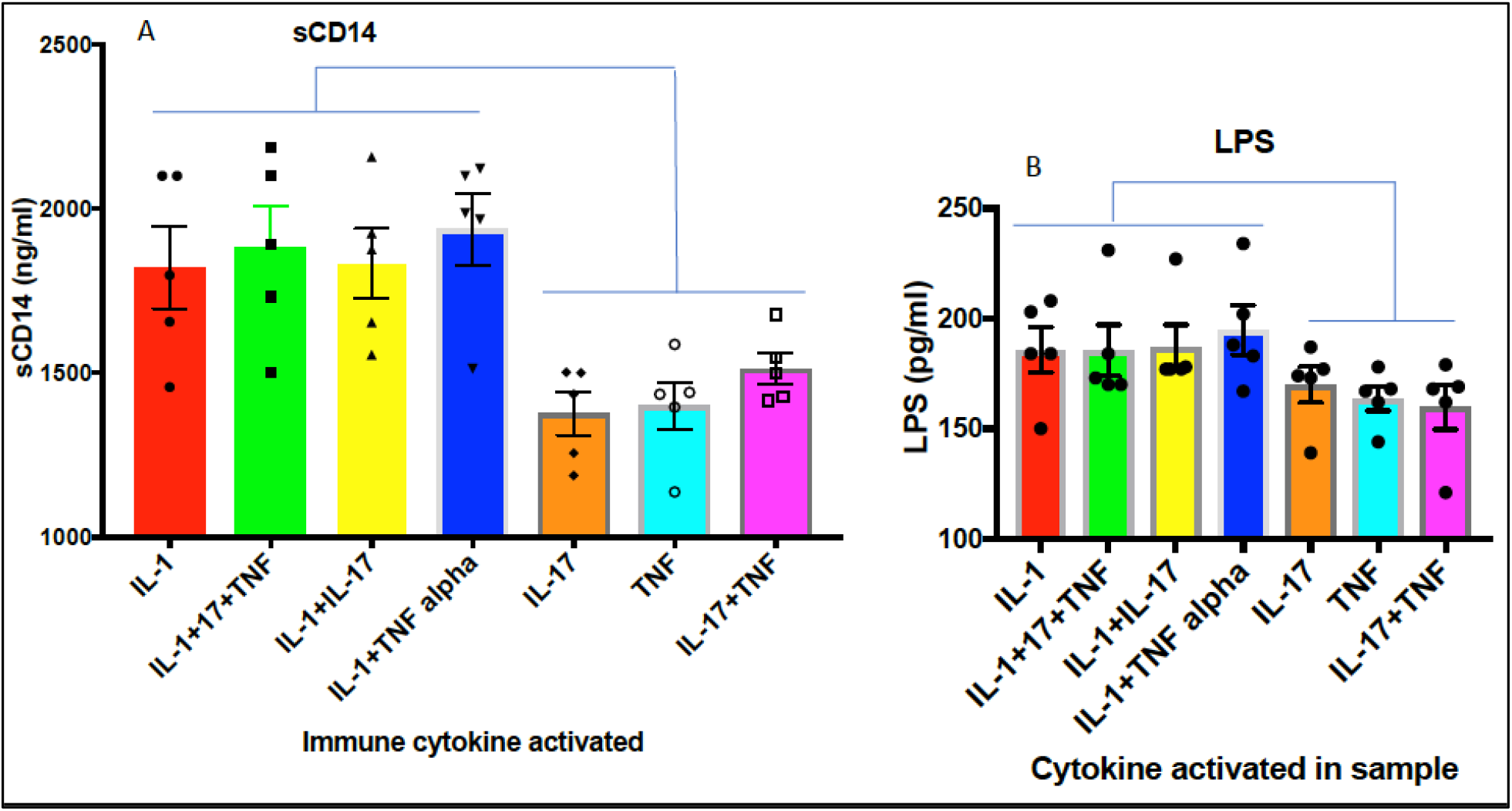

LPS and sCD14 were not significantly associated with the composite cognitive score at baseline (LPS: β=0.082/SD, 95%CI:-0.14, 0.31; sCD14: β=-0.031/SD, 95%CI:-0.21, 0.15).

Increased levels of LPS and sCD14 were significantly associated with cognitive decline. In adjusted models, LPS was directly associated with cognitive decline (adjusted β=-0.023 per SD/year, 95%CI:-0.036, −0.010). The association between sCD14 and cognitive decline was marginal (adjusted β=-0.018 per SD/year, 95%CI:-0.040, 0.004).

## Discussion

This pilot study provides evidence for the association between two ‘leaky gut’ markers, LPS and sCD14, and cognitive decline. Our observed elevated levels of sCD14 among individuals with high levels of IL-1 indicates a role of sCD14 in the IL-1 inflammatory pathway. This preliminary evidence suggests that gut microbiota-derived markers of gut permeability may drive systemic inflammation to levels that affect neuroinflammation and subsequent cognitive dysfunction.

Gut microbial dysbiosis and gut permeability (‘leaky gut’) increase the bioavailability of circulating microbial products that affect organs via activation of inflammatory signaling pathways. Lipopolysaccharide is an endotoxin produced by gram negative bacteria and a major driver of inflammation through activation of systemic monocytes and microglial cells, contributing to sustained neuroinflammation and neurodegeneration(11). High levels of circulatory sCD14 as a marker of monocyte activation are reflective of LPS exposure, although not strictly restricted to LPS activation(12). In the Framingham Heart and the Cardiovascular Health Study, higher levels of sCD14 were associated with a higher risk of dementia and brain aging MRI markers, and with decline in executive function(13). The highest sCD14 levels were associated with risk of dementia independent of vascular risk factors and circulating inflammatory markers such as CRP or IL-6, suggesting sCD14 as an important inflammatory marker of dementia.

IL-1 an inflammatory cytokine with diverse physiological functions and importance in innate immune processes(14). Small localized elevations in IL-1 may be sufficient to drive potent neuroinflammatory changes in the brain, affecting memory and other cognitive processes(15). The centrality of IL-1 elevations with increased sCD14 in plasma likely reflects shared upstream activators, such as LPS, involved in complex regulation through actions at toll-like receptors and at the NLRP3 inflammasome(15). Emerging understanding of IL-1’s function has identified possible roles in triggering innate and adaptive immune responses and their relative contributions to beneficial vs. detrimental outcomes in chronic vascular and neurodegenerative diseases. Our study suggests a possible link between IL-1 and gut dysbiosis with a potential paradigm shift in the understanding of balance between the beneficial and deleterious IL-1 functions.

A limitation is the small number of selected markers and participants. However, we purposely targeted our markers to gain preliminary data on the potential link between gut microbiota-derived markers and systemic inflammation that may lead to neuroinflammation and cognitive dysfunction. Since clinical studies on these markers are sparse, our preliminary results are valuable and promising. Larger targeted and untargeted gut microbiota-derived metabolomic studies are needed to better understand the ‘gut-microbiome-brain axis’, its role in neurovascular and neurocognitive dysfunction and to identify related targets for disease-modifying interventions.

## Data Availability

Anonymized data will be shared by request from any qualified investigator.

